# Generalizability and Clinical Implications of Electrocardiogram Denoising with Cardio-NAFNet

**DOI:** 10.1101/2022.10.26.22281565

**Authors:** Chanho Lim, Yunsung Chung, Jihun Hamm, Zhengming Ding, Mario Mekhael, Charbel Noujaim, Ala Assaf, Hadi Younes, Nour Chouman, Noor Makan, Eoin Donnellan, Nassir Marrouche

## Abstract

The rise of mobile electrocardiogram (ECG) devices came with the rise of frequent large magnitudes of noise in their recordings. Several artificial intelligence (AI) models have had great success in denoising, but the model’s generalizability and the enhancement in clinical interpretability are still questionable. We propose Cardio-NAFNet, a novel AI-based approach to ECG denoising by employing a modified version of Non-Linear Activation Free Network (NAFNET). We conducted three experiments for quantitative and qualitative evaluation of denoising, clinical implications and generalizability. In the first experiment, Cardio-NAFNet achieved 53.74dB average signal to noise ratio across varying magnitude of noise in beat-to-beat denoising, which is a significant improvement over the current state of the art model in ECG denoising. In the second experiment, we tested the enhancement in clinical interpretation of the ECG signals by utilizing a pretrained ECG classifier using 8 second long noise-free ECG signals. When the classifier was tested using noisy ECG signals and their denoised counterparts, Cardio-NAFNet’s denoised signals provided 26% boost in classification results. Lastly, we provide an external validation dataset composed of single-lead mobile ECG signals along with signal quality evaluation from physician experts. Our paper suggests a settling method to capture and reconstruct critical features of ECG signals not only in terms of quantitative evaluation, but also through generalizable qualitative evaluation.

## Introduction

With digital health evolution and numerous consumer electronics providing electrocardiograms (ECG), ECG denoising plays a pivotal role in standardizing and stabilizing the signals recorded amongst a multitude of devices and patients. Beyond providing a level of reliability of the mobile ECG recordings for physician’s interpretation, ECG denoising can play a critical role in translating the innovative artificial intelligence approaches using 12-lead ECG signals to the digital health realm. Previously, to reach beyond traditional use cases of electrocardiograms (ECG), numerous groups across the globe have provided methods to automate the processes typically done by subject matter experts and method to augment undiscovered knowledge about ECG signal’s discriminative features. For automated methods, a cardiologist-level arrhythmia detection and classification accuracy has been achieved using deep neural network [1]. Furthermore, the clinical implications of ECG signals has been expanded by an AI model detecting low ejection fraction using 12-lead ECG signals[2]. However, the AI models that were trained on clean 12-lead ECG in a hospital environment are bound to be inaccurate when tested with mobile ECG recorded during a patient or a consumer’s daily lives. Although measuring ECG signals has become more available to the public than ever, these recordings are frequently measured without any clinical staff’s oversight and more easily exposed to various types of noise. We learned throughout the years that ECG recordings are prone to three main types of noise - electrode motion (EM), baseline wandering (BW) and muscle artifacts (MA). Hence, effective methods to denoise ECG signals and experiments to evaluate its enhancements in clinical interpretability and generalizability in digital health realm are imperative.

ECG denoising methods can be largely divided into two categories - traditional denoising that relies on statistical methods and deep learning-based denoising models[3]. For example, traditional methods have seen success in ECG denoising using bandpass filters[4], empirical mode decomposition (EMD), Wavelet transformation methods[5, 6], adaptive filtering[7-10], and Bayesian filtering methods[11]. Simple bandpass filtering may be capable of rejecting low frequency noise like small baseline wandering and some high frequency noise such as jitters, but it often fails to cope with muscle artifacts and electrode motion artifacts that sporadically create false peaks and valleys. Kabir et al suggested an approach based on noise reduction algorithms in EMD and discrete wavelet transform domains, but the method is also limited to noise reduction with jitters and baseline wandering[12]. The recent advances in deep learning has impacted how ECG signals are processed, through new deep learning models such as autoencoders[13, 14], long short-term memory (LSTM)[15], generative adversarial network (GAN)[16, 17]. For example, Xiong et al utilized a combination of wavelet transform to deconstruct the signals and deep autoencoders (DAE)[18] to enhance the quality of corrupted signals. Others have also proposed stacked contractive denoising auto-encoder[19]. Both autoencoder based approaches were capable of removing BW, MA, EM and mixed noises at varying magnitudes. The generalizability of these models has been questioned by Wang et al., as autoencoder’s performances can be sensitive to its sample selection, which led them to suggest a GAN based method. Since introduced by Goodfellow et al in 2014, GAN variants have had remarkable contributions to the advancements of generative models. Pratik et al proposed a GAN framework that contains convolution layers in its generator and discriminator[20], but the model was only tested to prove its applications with individual types of noise, not any mixtures at varying magnitude. Xu et al utilized ResNet based GAN model but has demonstrated that that the model’s denoising capabilities diminished with larger noise samples at lower signal to noise ratio[17]. Wang et al proposed a conditional generative adversarial network (CAE-CGAN) framework where they utilize a convolutional U-Net architecture as a generator, a discriminator with least squared loss, and a pretrained support vector machine (SVM) based classifier that learns to classify each beat[21]. Upon our review, CAE-CGAN’s methods were deemed the most sound as it demonstrated promising improvement in SNR across individual and mixture of noise at varying SNR while also proving that the denoised signal also enhances classification accuracy for each denoised beat.

We note that the majority of the denoising work has been done by combining the noise from MIT-BIH noise stress database with clean ECG signals from various ECG databases in Physionet’s MIT-BIH Databases[22, 23], specifically the Arrhythmia Database. Despite numerous authors highlighting the rise of wearables and other mobile devices for ECG recordings as one of the primary motivations for denoising, the majority of the models are only evaluated internally within the arrhythmia database that was collected during 1970s. Also, most of the models prioritize on the quantitative evaluation of denoising using signal to noise ratio (SNR), but the qualitative evaluation of the signals is often missing as only a few have performed tests to confirm that denoising also improves clinical interpretability.

To address these issues, we propose Cardio-NAFNet, a non-linear activation free network for ECG denoising. Cardio-NAFNet utilizes the current (SOTA) framework used in image denoising domain with reduced dimensionality and complexity along with separate loss functions to tailor the framework towards ECG signal denoising. We conducted three experiments designed to independently prove Cardio-NAFNet’s superior performance to the current SOTA model in an identical environment using the arrhythmia database, enhanced clinical interpretability through rhythm-based classification, and generalizability with an external validation dataset composed of real-world mobile ECG signals.

## Method

### Experiment Design

The first experiment’s objective is to evaluate Cardio-Net’s performance against that of the current SOTA model (CAE-CGAN)[21] in an identical testing environment. We prepared 10 records from Physionet’s arrhythmia database[24]. The 10 records are 100, 101, 106, 112, 117, 121, 123, 209, 220, and 228 and uses MLII lead. Then we split each record into samples with lengths of 512, which is about 1.2 seconds with the dataset’s sampling rate of 360Hz. For training and testing, we used 8:2 random split.

The second experiment’s objective is to validate our argument that denoised samples should not only have enhanced SNR, but also improved interpretability. We aim to demonstrate improved classification results with beat and rhythm labels. The records were resampled to 64Hz, then split into samples length of 512, which is 8 seconds long. To split the signals, we visited every annotation point, which exists with every beat, then chose point at random to be the center of the sample, where the distance from the center to the annotation point was always less than the quarter of the total sample length. With this method, we were able to create samples that were multi-labeled with their rhythm types and their beat types. We trained a convolution neural network (CNN) classifier with clean ECG samples from the arrhythmia database, then evaluated its performance using unseen clean samples, noisy samples, and denoised samples.

The third experiment was designed to highlight the generalizability of our model by utilizing an independent dataset from DECAAF-II[25]. We retrained Cardio-NAFNet to suit the samples from DECAAF-II[25], which are measured at 200Hz with a 20 second window, providing sample length of 4000. The training data was generated using the same framework as the second experiment, but with sampling rate of 200Hz and sample length of 20 seconds. After the samples were denoised, we handed the samples over to the expert reviewers at Tulane University’s Heart and Vascular Institute.

### Study Data

The internal training and validation data are from two databases on Physionet. We pulled the ECG recordings from MIT-BIH Arrhythmia Database[24], and the three different types of noises from MIT-BIH Noise Stress Database[26]. The MIT-BIH Arrhythmia database is from 4000 long-term Holter recordings that were obtained from Beth Israel Hospital Arrhythmia Laboratory. The arrhythmia database contains 23 records that were chosen at random from the aforementioned dataset, and 25 recordings that were selected for containing clinically important phenomena. Overall, the database contains 48 records where the average length of the records is around 30 minutes long. While most records have modified limb lead II (MLII) as the first lead, a few records did not contain MLII due to surgical dressings on the patients, hence we removed records 102, and 104 from the dataset. All recordings are digitized at a sampling rate of 360Hz. The recordings in the database are labeled with 20 categories of beat annotations and 15 categories of rhythm annotations. The subjects were 25 men aged 32 to 89 years, and 22 women aged 23 to 89 years.

The MIT-BIH Noise Stress Database includes three half hour recordings of 3 types of noise typical in ambulatory ECG recordings. The three noise records are baseline wander (BW), muscle artifact (MA), and electrode motion (EM) artifact. To evaluate the denoising capabilities of our model in comparison with the results in CAE-CGAN[21], we created 42 different scenarios of denoising which are combinations of the three noise types (EM, BW, MA, EM+BW, MA+BW, EM+MA, EM+MA+BW) and varying levels of signal to noise ratio (SNR) from 0dB to 5dB.

An external validation dataset was prepared to ensure the generalizability of Cardio-NAFNet. We randomly selected 222 ECG strips from the DECAAF-II Trial[25], which are the recordings used to track the outcome of 843 patients who received atrial fibrillation ablation from 44 sites around the world. The strips are recorded using single-lead handheld devices called “ECG Check”. The length of the recordings are generally around 30 seconds with a sampling rate of 200Hz. As the strips are unfiltered raw recordings from a handheld device, we deem the recordings here to be “real world” examples of noisy ECG signals with large variance in noise types and magnitude. The strips were thoroughly reviewed by intra and inter reviewers that were all expert physicians.

### Preprocessing

As the objective of the three experiments differ, the length of the samples in each experiment also differs. In the first experiment, we pulled record 103, 105, 111, 116, 122, 205, 213, 219, 223, 230 for training and sliced the records to sample lengths of 512. Considering the sampling rate of 360Hz in the arrhythmia database, each input signals are roughly 1.4 seconds long. For the second experiment, we wanted to preserve the rhythm labels; hence, we resampled the records to 64Hz, then the sample lengths of 512 again, resulting with 8 second strips. For the final experiment, we resampled the signals to 200Hz to match the sampling rate of the records in the external validation dataset, then sliced the records to sample lengths of 4000, resulting with 20 second strips. After resampling and slicing, all training and internal validation samples went through the steps below to generate simulated noisy signals.

The generation of the training data is intuitive. We inject the combinations of noise into the clean ECG samples from the arrhythmia database, arriving at three different variations of the signals – the clean ECG samples, the injected noise, and the simulated noisy ECG sample. The objective of the Cardio-NAFNet is to receive simulated noisy ECG samples and generate denoised samples that closely resemble their corresponding original ECG samples. When injecting the noise into the original ECG signals, we measure the signal to noise ratio (SNR) by the following equation.

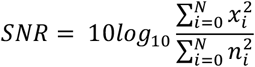

To provide various mixtures of noise by providing a random length, a random signal to noise ratio (SNR) to a randomized segment in an ECG signal. For validating samples, we created 42 different testing environments by fixing the signal to ratio to integers from 0dB to 5dB and providing all combinations of baseline wander, muscle artifacts, and electrode motion artifacts. We fixed the signal to ratio of generated noisy signals by calculating a that is provided by the following equation:

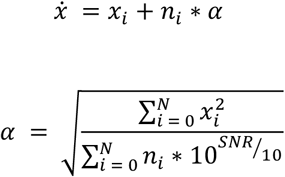

where 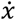 represents the simulated noisy signals, *x* represents individual clean ECG sample from the arrhythmia database, *n* is the noise, *S* is the number of samples, and *a* represents the constant that is multiplied to the noise to generate noisy samples at fixed SNR. With the formulas above, we generate combinations of simulated noisy ECG samples at fixed SNR from 0dB to 5dB with all combinations of noise types. We then normalized the signals using min-max normalization:

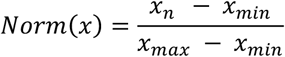

For the external validation dataset, we chose to slice the recordings into 20 second windows by choosing the starting point of the window to be a random point in the first 10 seconds of the signal due to a small variance in the length of the recordings. We also performed min-max normalization to all samples.

### Network Architecture

Our Cardio-NAFNet resembles the original structure of NAFNet[27] with reduced complexity and dimension to transform the model’s original framework dedicated to 2-dimentional image restoration to ECG signal restoration. The network follows U-Net architecture where we utilize an encoder and a decoder with skip connections. The encoder is comprised of 10 NAFBlocks and the decoder is comprised of 4 NAFBlocks as shown in Figure 1b. The generalizability of encoder-decoder architecture has been questioned before, and we provide evidence that model performance holds with an external validation dataset.

**Figure 1:**
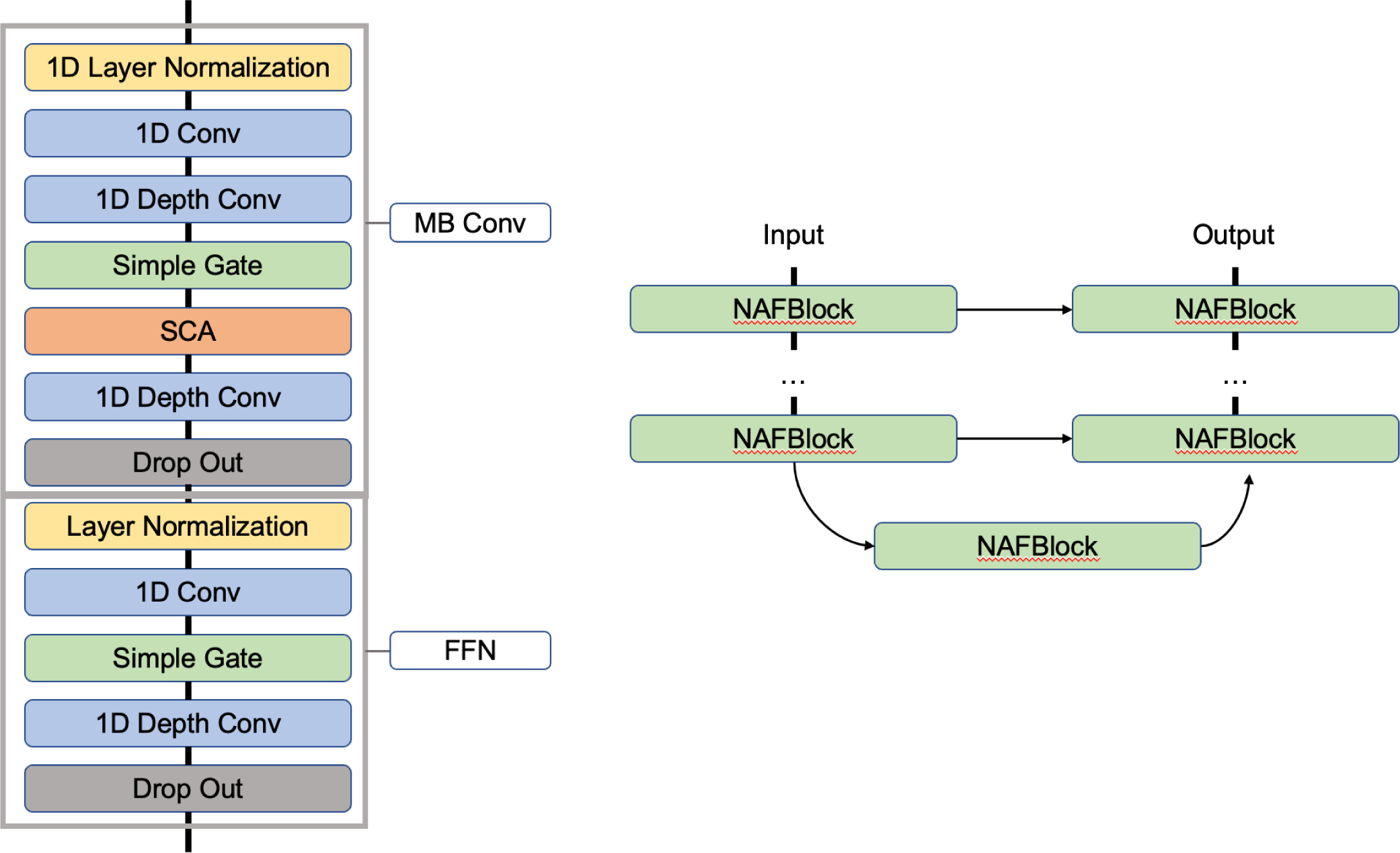
Figure 1a describes the structure of the Block using Mobile Convolution (MB Block) and Feed Forward Network (FFN) Block with changes to the attention layers using simple channel attention (SCA) and simple gate along with drop out layers. Figure 2b describes the overall U-shaped architecture of Cardio-NAFNet where the left side represents encoder with 10 NAFBlocks and the right side is the decoder with 4 NAFBlocks. The decoder also receives maps from the encoder blocks using skip connection.

For training, Cardio-NAFNet receives batches that comprise pairs of noisy ECG signal generated from the preprocessing steps and their corresponding original ECG strips unaltered by noise. The matching original ECG signals are only used to calculate the loss by taking the distance of the denoised output to the original signal.

Each NafNet’s Block consists of layers without nonlinear activation functions (e.g., sigmoid, softmax, ReLu, etc). The block consists of Layer Normalization, pointwise convolution, depth wise convolution, simple channel attention, simple gate, elementwise multiplication/addition, and dropout layers in the order described in figure. The core difference between NAFNet’s Block versus the feed forward networks (FFN) in transformers is in the simple gate, which allows the entire block to be free of nonlinear activation functions. See Figure 1a for the structure of NAFBlock.

Mean Squared Error (MSE) *L*_*MSE*_ is adopted to measure the differences between denoised signals and clean signals. Similar to Wang et al., *L*_*max*_ is used to measure the maximum difference between denoised and clean signals. It helps the model to capture the local characteristics of ECG signals.

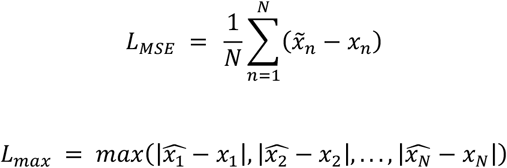

where 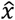 indicates denoised signals and *x* indicates clean signals. N represents the total number of samples. Our total loss function is defined as:

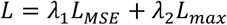

where *λ*_1_ and *λ*_2_ are weighted coefficients. Through our experiments, we chose *λ*_1_ = 0.8 and *λ*_2_ = 0.2. We train models with AdamW optimizer with learning rate of 0.0001 (*β*_1_ = 0.9, *β*_2_ = 0.999). The batch size is 256.

### Evaluation

The performance is measured by root mean square error (RMSE) and SNR as follows:

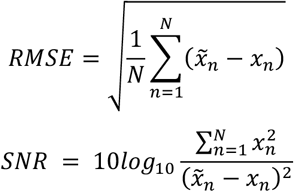

where *x* is the original clean signal, 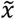 is denoised signal, and *S* is the number of samples. RMSE indicates the difference between two signals. While the SNR formula for the evaluation may seem different from the one introduced to prepare the training samples, both formulas essentially represent the same ratio of the ECG signal to the noise as 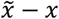 is the remaining noise after the signal was denoised. The RMSE and SNR possess an inverse relationship where smaller RMSE values indicate larger SNR. Cardio-NAFNet’s objective is to minimize RMSE and maximize SNR, which indicates a stronger power of the ECG signal to the noise.

The samples from DECAAF-II dataset were only used for external validation. We highlight that the samples from DECAAF-II dataset are real world examples of unfiltered mobile ECG samples as the patients submitted the data from home during the follow up period of the trial; thus, it is impossible to measure the SNR of these samples as we do not have a clean version, nor the noise separated from the signal. We provided 222 original samples and their corresponding denoised samples to the physicians at Tulane University’s Heart and Vascular institute to review the quality of denoising with the following scale.

#### Signal Quality Scale

1. Uninterpretable
2. Signal suffers from heavy combinations of baseline wandering, muscle artifacts and etc. Some beats are not recoverable, but the trend of the rhythm is identifiable to make an educated guess
3. Signal demonstrates heavy amplitudes of noise, but all beats are clear and rhythm is identifiable
4. Signal contains very minor noise but the rhythm is interpretable
5. Signal shows no presence of noise

## Results

In the first experiment, we created an identical environment to that of CAE-CGAN’s experiment to provide a direct comparison of Cardio-NAFNet’s performance to CAE-CGAN’s performance[21]. Table 1 demonstrates that Cardio-NAFNet’s performance has a significant improvement in all noise combinations at all noise levels, resulting in a combined average difference of 11.76dB. In the supplement, we also provide results to compare the results with not only CGAN, but also with Improved denoising autoencoder[13], and adversarial method[21, 28, 29]. We note that while our model follows the general autoencoder architecture, the skip connections from the encoder to the decoder and utilizing NAFBlocks instead of ConvBlocks provide a significant improvement in results.

**Table 1:** Denoising results of 360Hz sampling rate by noise type and SNR

| SNR | Methods | Denoised Metrics | Noise Type |  |  |  |  |  |  |  |
| --- | --- | --- | --- | --- | --- | --- | --- | --- | --- | --- |
|  |  |  | BW | EM | MA | BW+EM | BW+MA | MA+EM | BW+MA+EM | Avg. |
| 0dB | CGAN | SNR(dB) | 39.59 | 40.09 | 39.09 | 40.08 | 40.43 | 40.38 | 39.49 | 39.88 |
|  |  | RMSE | 0.0031 | 0.0029 | 0.0033 | 0.0029 | 0.0028 | 0.0029 | 0.0031 | 0.0030 |
|  | Cardio-NAFNET (proposed) | SNR(dB) | 53.38 | 52.97 | 51.70 | 51.80 | 52.19 | 51.31 | 51.55 | 52.13 |
|  |  | RMSE | 0.0027 | 0.0014 | 0.0024 | 0.0018 | 0.0026 | 0.0016 | 0.0017 | 0.0020 |
| 1dB | CGAN | SNR(dB) | 41.29 | 42.31 | 41.21 | 42.42 | 42.57 | 42.67 | 41.92 | 42.06 |
|  |  | RMSE | 0.0025 | 0.0022 | 0.0025 | 0.0022 | 0.0021 | 0.0022 | 0.0023 | 0.0023 |
|  | Cardio-NAFNET (proposed) | SNR(dB) | 54.69 | 54.45 | 53.27 | 53.05 | 53.75 | 52.75 | 53.04 | 53.57 |
|  |  | RMSE | 0.0016 | 0.0010 | 0.0018 | 0.0013 | 0.0020 | 0.0013 | 0.0013 | 0.0015 |
| 2dB | CGAN | SNR(dB) | 41.87 | 43.04 | 41.78 | 43.22 | 43.23 | 43.38 | 42.66 | 42.77 |
|  |  | RMSE | 0.0023 | 0.0020 | 0.0023 | 0.0020 | 0.0020 | 0.0020 | 0.0021 | 0.0021 |
|  | Cardio-NAFNET (proposed) | SNR(dB) | 55.39 | 54.89 | 53.89 | 53.54 | 54.52 | 53.28 | 53.66 | 54.17 |
|  |  | RMSE | 0.0013 | 0.0010 | 0.0014 | 0.0012 | 0.0015 | 0.0013 | 0.0011 | 0.0013 |
| 3dB | CGAN | SNR(dB) | 42.09 | 43.26 | 41.96 | 43.41 | 43.36 | 43.65 | 42.86 | 42.94 |
|  |  | RMSE | 0.0023 | 0.0020 | 0.0023 | 0.0019 | 0.0020 | 0.0019 | 0.0021 | 0.0021 |
|  | Cardio-NAFNET (proposed) | SNR(dB) | 55.86 | 55.07 | 54.10 | 53.72 | 54.99 | 53.47 | 53.96 | 54.45 |
|  |  | RMSE | 0.0010 | 0.0009 | 0.0013 | 0.0011 | 0.0012 | 0.0013 | 0.0010 | 0.0011 |
| 4dB | CGAN | SNR(dB) | 41.96 | 43.04 | 41.74 | 43.20 | 43.13 | 43.40 | 42.65 | 42.73 |
|  |  | RMSE | 0.0023 | 0.0021 | 0.0024 | 0.0020 | 0.0020 | 0.0020 | 0.0021 | 0.0021 |
|  | Cardio-NAFNET (proposed) | SNR(dB) | 55.92 | 54.95 | 54.04 | 53.63 | 55.09 | 53.37 | 53.87 | 54.41 |
|  |  | RMSE | 0.0012 | 0.0010 | 0.0021 | 0.0014 | 0.0017 | 0.0014 | 0.0012 | 0.0014 |
| 5dB | CGAN | SNR(dB) | 41.10 | 41.75 | 40.59 | 41.80 | 41.93 | 42.03 | 41.28 | 41.50 |
|  |  | RMSE | 0.0026 | 0.0024 | 0.0027 | 0.0024 | 0.0023 | 0.0023 | 0.0025 | 0.0025 |
|  | Cardio-NAFNET (proposed) | SNR(dB) | 55.52 | 54.26 | 53.14 | 53.04 | 54.41 | 52.56 | 53.17 | 53.73 |
|  |  | RMSE | 0.0024 | 0.0011 | 0.0033 | 0.0017 | 0.0021 | 0.0016 | 0.0014 | 0.0019 |

**Table 2:** Comparison of EM denoising results.

| Methods | Input SNR | Denoised Metrics | Record Number |  |  |  |  |  |  |  |  |  |  |
| --- | --- | --- | --- | --- | --- | --- | --- | --- | --- | --- | --- | --- | --- |
|  |  |  | 103 | 105 | 111 | 116 | 122 | 205 | 213 | 219 | 223 | 230 | Avg. |
| Improved DAE | 0dB | SNR(dB) | 22.75 | 23.70 | 23.39 | 21.34 | 17.70 | 23.47 | 19.33 | 18.38 | 23.17 | 22.40 | 21.56 |
|  |  | RMSE | 0.0290 | 0.0330 | 0.0340 | 0.0350 | 0.0500 | 0.0330 | 0.0400 | 0.0410 | 0.0310 | 0.0390 | 0.0365 |
| Adversarial Method |  | SNR(dB) | 38.09 | 34.27 | 33.07 | 30.02 | 28.74 | 38.44 | 30.27 | 28.24 | 31.75 | 30.87 | 32.38 |
|  |  | RMSE | 0.0050 | 0.0080 | 0.0093 | 0.0115 | 0.0134 | 0.0048 | 0.0125 | 0.0150 | 0.0101 | 0.0119 | 0.0102 |
| CGAN |  | SNR(dB) | 39.49 | 38.89 | 39.65 | 40.97 | 39.76 | 38.45 | 41.28 | 40.40 | 39.72 | 42.34 | 40.09 |
|  |  | RMSE | 0.0022 | 0.0032 | 0.0040 | 0.0027 | 0.0026 | 0.0026 | 0.0031 | 0.0027 | 0.0029 | 0.0034 | 0.0029 |
| Cardio-NAFNET (proposed) |  | SNR(dB) | 51.73 | 47.30 | 45.46 | 46.97 | 49.21 | 53.04 | 46.09 | 47.95 | 49.16 | 49.82 | 48.67 |
|  |  | RMSE | 0.0022 | 0.0036 | 0.0058 | 0.0045 | 0.0039 | 0.0017 | 0.0054 | 0.0042 | 0.0061 | 0.0036 | 0.0041 |
| Improved DAE | 1.25dB | SNR(dB) | 22.97 | 23.94 | 23.57 | 21.82 | 18.76 | 23.57 | 19.79 | 19.07 | 23.55 | 22.54 | 21.96 |
|  |  | RMSE | 0.0290 | 0.0330 | 0.0330 | 0.0330 | 0.0420 | 0.0330 | 0.0370 | 0.0380 | 0.0300 | 0.0380 | 0.0346 |
| Adversarial Method |  | SNR(dB) | 38.56 | 34.79 | 33.45 | 30.77 | 29.28 | 38.96 | 30.68 | 29.21 | 32.19 | 31.11 | 32.90 |
|  |  | RMSE | 0.0049 | 0.0075 | 0.0089 | 0.0105 | 0.0126 | 0.0046 | 0.0119 | 0.0134 | 0.0096 | 0.0116 | 0.0096 |
| CGAN |  | SNR(dB) | 42.34 | 42.26 | 42.75 | 44.20 | 43.00 | 41.46 | 44.93 | 43.69 | 42.74 | 45.48 | 43.28 |
|  |  | RMSE | 0.0016 | 0.0021 | 0.0027 | 0.0018 | 0.0018 | 0.0017 | 0.0020 | 0.0018 | 0.0020 | 0.0023 | 0.0020 |
| Cardio-NAFNET (proposed) |  | SNR(dB) | 53.32 | 49.05 | 48.91 | 49.03 | 51.06 | 54.98 | 47.31 | 49.31 | 51.15 | 52.57 | 50.67 |
|  |  | RMSE | 0.0016 | 0.0030 | 0.0059 | 0.0025 | 0.0021 | 0.0013 | 0.0053 | 0.0037 | 0.0034 | 0.0036 | 0.0032 |
| Improved DAE | 5dB | SNR(dB) | 23.45 | 24.66 | 23.65 | 23.08 | 20.81 | 23.66 | 20.69 | 21.01 | 24.00 | 22.81 | 22.78 |
|  |  | RMSE | 0.0270 | 0.0300 | 0.0330 | 0.0300 | 0.0350 | 0.0300 | 0.0340 | 0.0300 | 0.0280 | 0.0370 | 0.0314 |
| Adversarial Method |  | SNR(dB) | 39.39 | 35.67 | 34.10 | 21.72 | 30.01 | 39.89 | 31.37 | 31.23 | 32.96 | 31.53 | 32.79 |
|  |  | RMSE | 0.0044 | 0.0068 | 0.0082 | 0.0095 | 0.0116 | 0.0041 | 0.0110 | 0.0106 | 0.0088 | 0.0111 | 0.0086 |
| CGAN |  | SNR(dB) | 41.05 | 40.72 | 41.18 | 42.65 | 41.89 | 40.29 | 42.72 | 42.03 | 41.43 | 43.56 | 41.75 |
|  |  | RMSE | 0.0018 | 0.0033 | 0.0056 | 0.0022 | 0.0020 | 0.0021 | 0.0026 | 0.0022 | 0.0024 | 0.0029 | 0.0027 |
| Cardio-NAFNET (proposed) |  | SNR(dB) | 54.31 | 50.01 | 51.37 | 50.20 | 52.12 | 55.79 | 49.42 | 51.08 | 52.45 | 52.93 | 51.97 |
|  |  | RMSE | 0.0010 | 0.0028 | 0.0016 | 0.0015 | 0.0011 | 0.0008 | 0.0023 | 0.0023 | 0.0013 | 0.0012 | 0.0015 |

For the second experiment, we demonstrate that not only our model performance holds when we stretch the input to 8 seconds, but also the ECG rhythm classifications drastically improved after denoising. The detailed SNR and RMSE results with 8 second samples can be found in Table 5 and 6. Figure 2 shows the original clean ECG from the arrhythmia database, a simulated ECG through our preprocessing, and the output of Cardio-NAFNet when it receives the simulated ECGs. As shown in the figure, while the simulated noisy signals contain a significant amount of noise, the denoised samples are nearly indistinguishable from the clean ECG signals during validation. Also, a pretrained 4 label classifier that achieved .98 F-1 score with clean signals from the arrhythmia database was applied to the noisy signals and corresponding denoised signals. When applied with different noise types demonstrated in Table7, denoised signals had 26% average improvement compared to the noisy signals.

**Figure 2:**
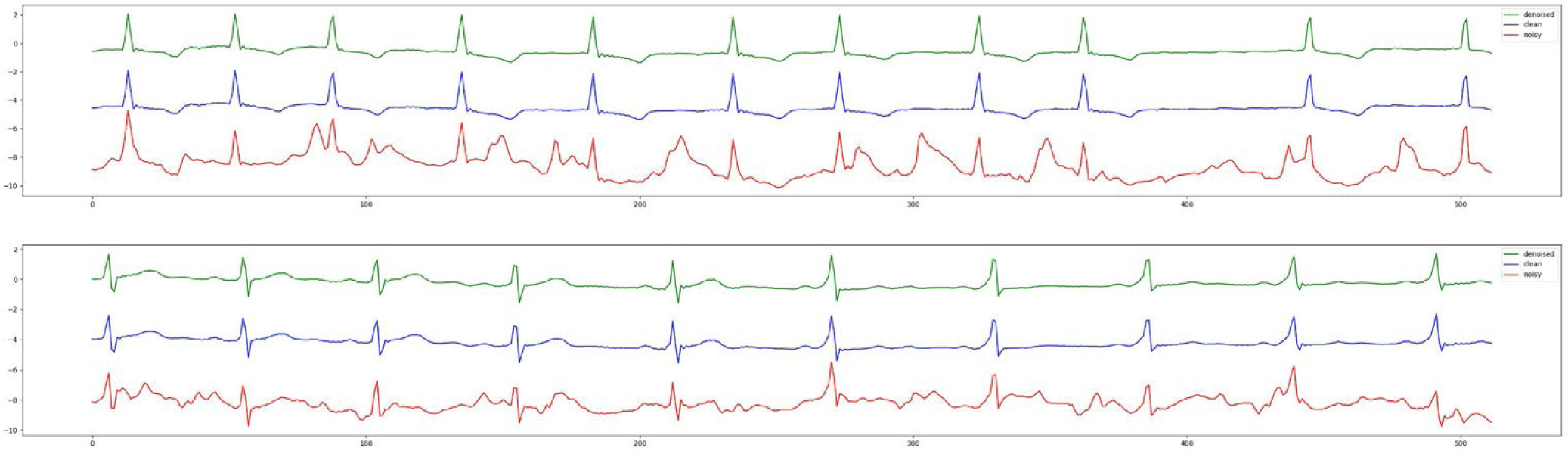
The denoised outputs from Cardio-NAFNet are in green, the clean ECG signals from the arrhythmia database are in blue, and the simulated noisy signals are in red. Cardio-NAFNet takes the signals in red as an input and uses the signals in blue only to calculate the loss to produce the signals in green. The denoised outputs here are nearly indistinguishable from the original clean ECG samples.

For the third experiment, we highlight the generalizability of Cardio-NAFNet’s by providing the denoised results from single-lead mobile ECG samples. The original samples and corresponding denoised examples can be found in Figure 3. Above visual representation, the denoised signals were reviewed by expert physicians with the metric provided in Evaluation. The improved results can be found in Table 3. In our proposed metric scaling from 1 to 5, the expert physicians’ scored the unfiltered signals from the DECAAF-II a mean of 3.18 with a variance of 0.94, while the denoised signals achieved 4.46 with a variance of 0.91. We noticed that majority of the unfiltered recordings that were in 3 or 4 range, meaning individual beats were identifiable, but with the presence of noise, was scored 5 after denoising.

**Figure 3:**
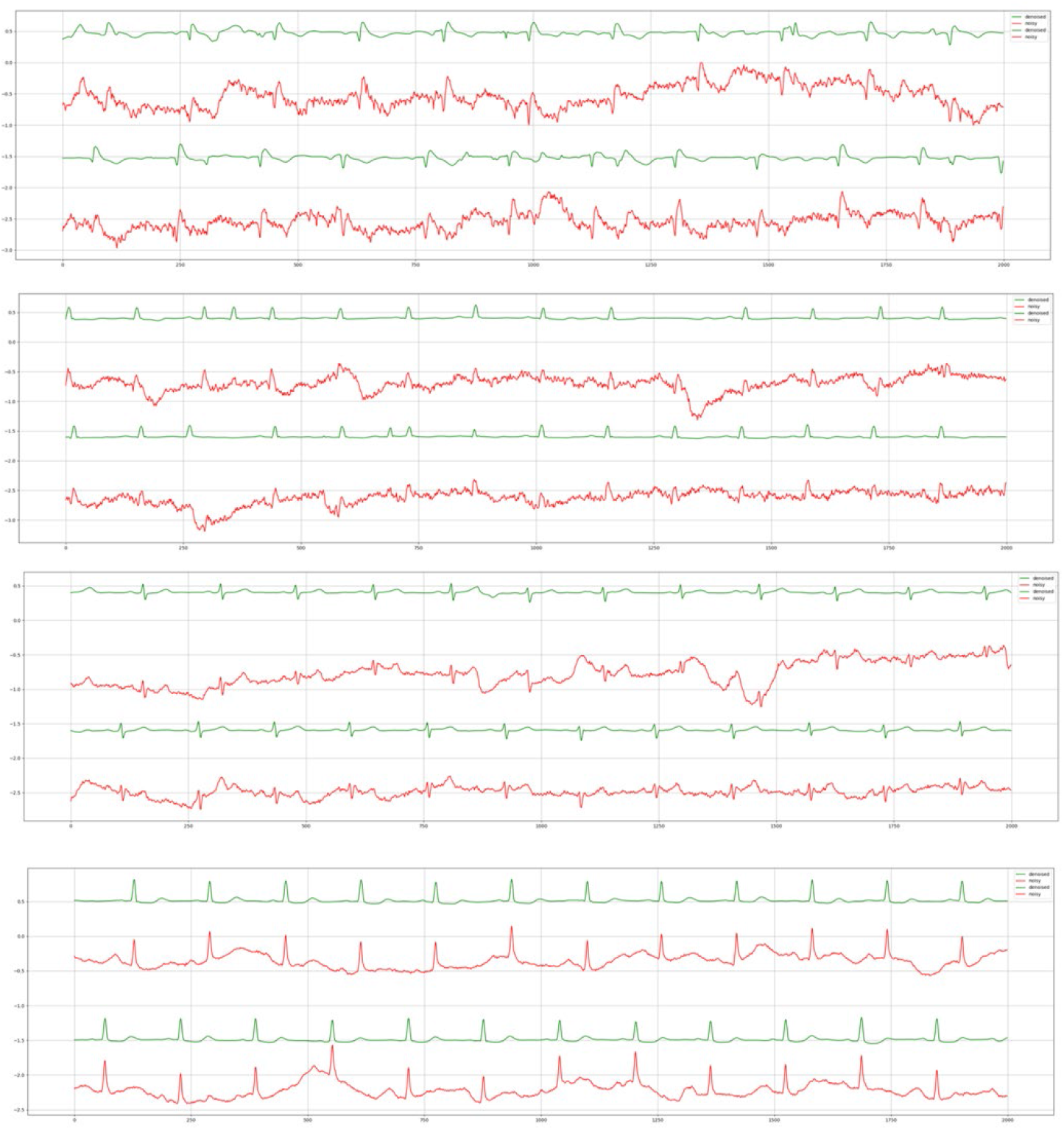
The denoised outputs from Cardio-NAFNet are in green and the unfiltered mobile ECG signals from DECAAF-II trial are in red. The records are in an order by their original signal quality score ranging from one to four.

**Table3:**
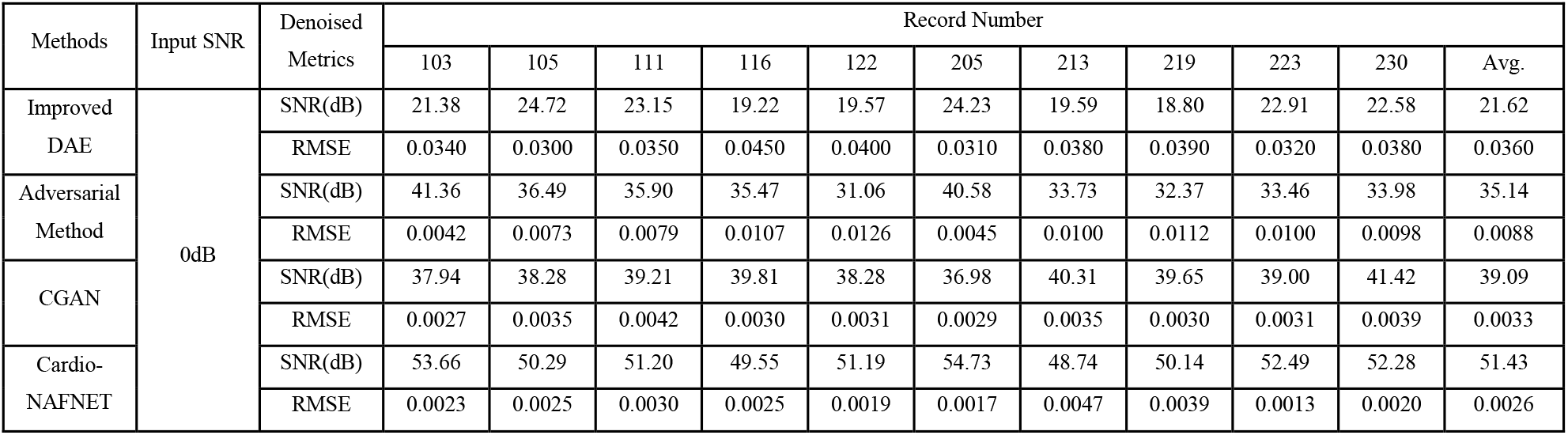
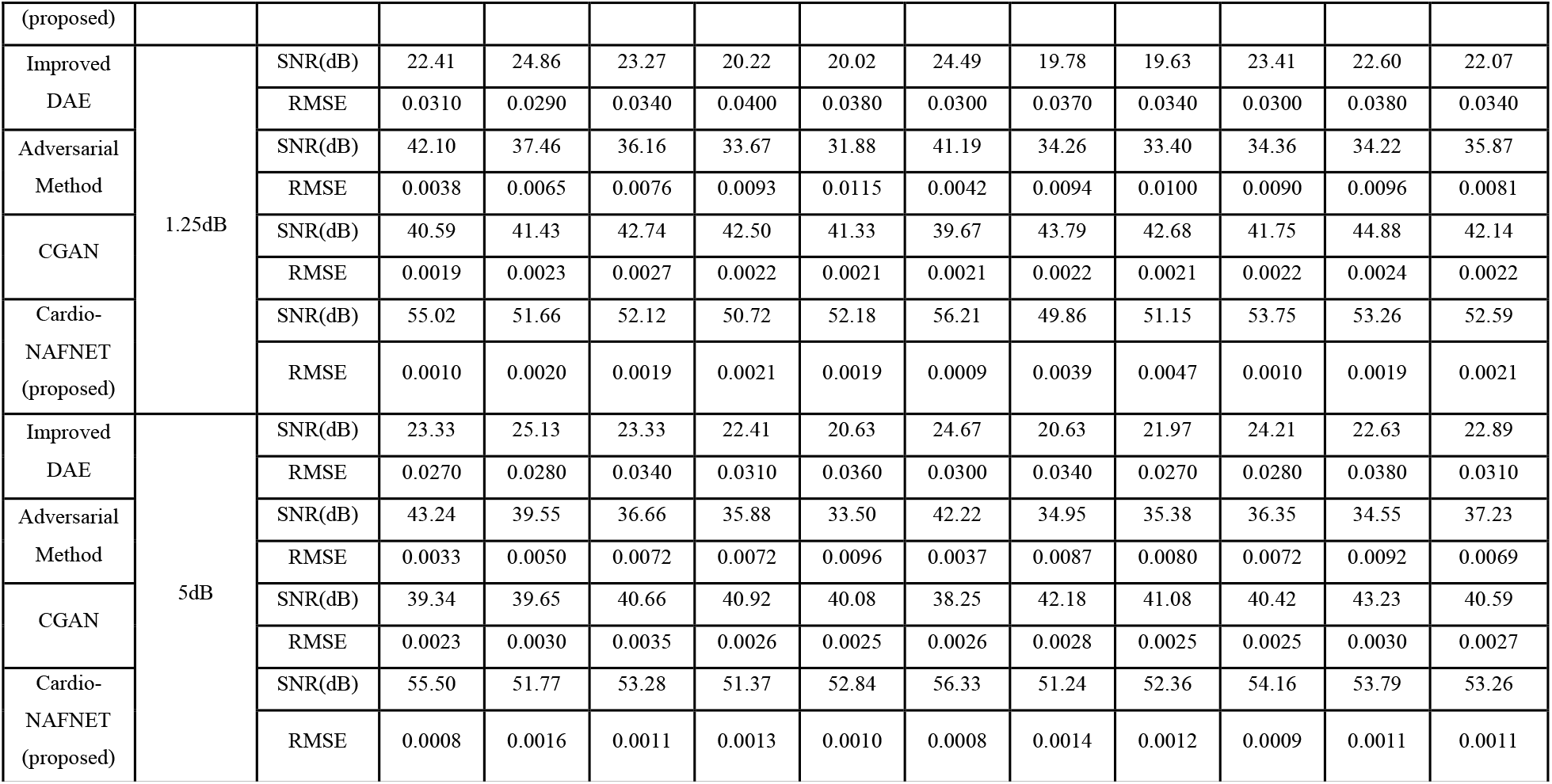
Comparison of MA denoising results.

**Table4:**
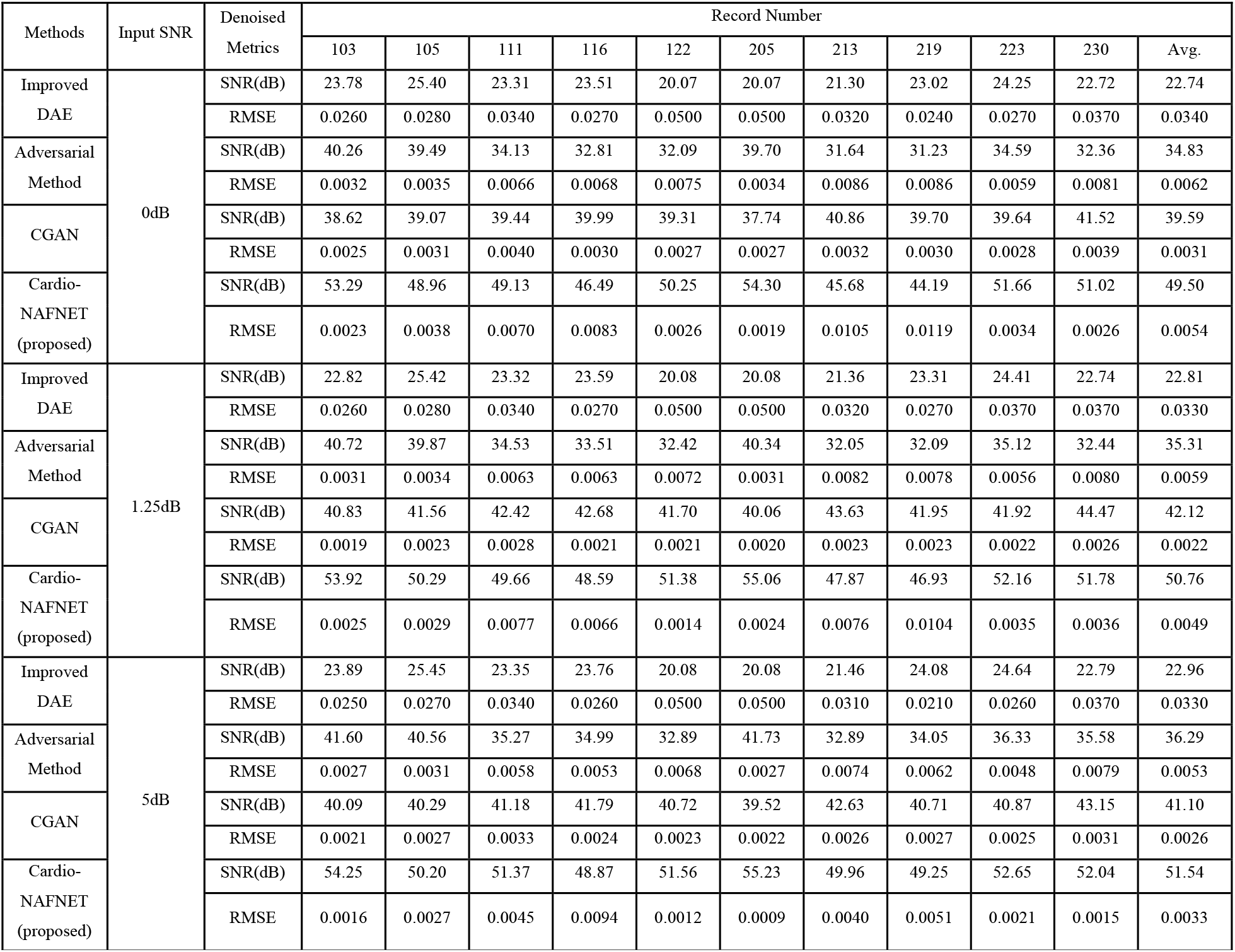
Comparison of BW denoising results.

| Methods | Input SNR | Denoised Metrics | Record Number |  |  |  |  |  |  |  |  |  |  |
| --- | --- | --- | --- | --- | --- | --- | --- | --- | --- | --- | --- | --- | --- |
|  |  |  | 103 | 105 | 111 | 116 | 122 | 205 | 213 | 219 | 223 | 230 | Avg. |
| Improved DAE | 0dB | SNR(dB) | 23.78 | 25.40 | 23.31 | 23.51 | 20.07 | 20.07 | 21.30 | 23.02 | 24.25 | 22.72 | 22.74 |
|  |  | RMSE | 0.0260 | 0.0280 | 0.0340 | 0.0270 | 0.0500 | 0.0500 | 0.0320 | 0.0240 | 0.0270 | 0.0370 | 0.0340 |
| Adversarial Method |  | SNR(dB) | 40.26 | 39.49 | 34.13 | 32.81 | 32.09 | 39.70 | 31.64 | 31.23 | 34.59 | 32.36 | 34.83 |
|  |  | RMSE | 0.0032 | 0.0035 | 0.0066 | 0.0068 | 0.0075 | 0.0034 | 0.0086 | 0.0086 | 0.0059 | 0.0081 | 0.0062 |
| CGAN |  | SNR(dB) | 38.62 | 39.07 | 39.44 | 39.99 | 39.31 | 37.74 | 40.86 | 39.70 | 39.64 | 41.52 | 39.59 |
|  |  | RMSE | 0.0025 | 0.0031 | 0.0040 | 0.0030 | 0.0027 | 0.0027 | 0.0032 | 0.0030 | 0.0028 | 0.0039 | 0.0031 |
| Cardio-NAFNET (proposed) |  | SNR(dB) | 53.29 | 48.96 | 49.13 | 46.49 | 50.25 | 54.30 | 45.68 | 44.19 | 51.66 | 51.02 | 49.50 |
|  |  | RMSE | 0.0023 | 0.0038 | 0.0070 | 0.0083 | 0.0026 | 0.0019 | 0.0105 | 0.0119 | 0.0034 | 0.0026 | 0.0054 |
| Improved DAE | 1.25dB | SNR(dB) | 22.82 | 25.42 | 23.32 | 23.59 | 20.08 | 20.08 | 21.36 | 23.31 | 24.41 | 22.74 | 22.81 |
|  |  | RMSE | 0.0260 | 0.0280 | 0.0340 | 0.0270 | 0.0500 | 0.0500 | 0.0320 | 0.0270 | 0.0370 | 0.0370 | 0.0330 |
| Adversarial Method |  | SNR(dB) | 40.72 | 39.87 | 34.53 | 33.51 | 32.42 | 40.34 | 32.05 | 32.09 | 35.12 | 32.44 | 35.31 |
|  |  | RMSE | 0.0031 | 0.0034 | 0.0063 | 0.0063 | 0.0072 | 0.0031 | 0.0082 | 0.0078 | 0.0056 | 0.0080 | 0.0059 |
| CGAN |  | SNR(dB) | 40.83 | 41.56 | 42.42 | 42.68 | 41.70 | 40.06 | 43.63 | 41.95 | 41.92 | 44.47 | 42.12 |
|  |  | RMSE | 0.0019 | 0.0023 | 0.0028 | 0.0021 | 0.0021 | 0.0020 | 0.0023 | 0.0023 | 0.0022 | 0.0026 | 0.0022 |
| Cardio-NAFNET (proposed) |  | SNR(dB) | 53.92 | 50.29 | 49.66 | 48.59 | 51.38 | 55.06 | 47.87 | 46.93 | 52.16 | 51.78 | 50.76 |
|  |  | RMSE | 0.0025 | 0.0029 | 0.0077 | 0.0066 | 0.0014 | 0.0024 | 0.0076 | 0.0104 | 0.0035 | 0.0036 | 0.0049 |
| Improved DAE | 5dB | SNR(dB) | 23.89 | 25.45 | 23.35 | 23.76 | 20.08 | 20.08 | 21.46 | 24.08 | 24.64 | 22.79 | 22.96 |
|  |  | RMSE | 0.0250 | 0.0270 | 0.0340 | 0.0260 | 0.0500 | 0.0500 | 0.0310 | 0.0210 | 0.0260 | 0.0370 | 0.0330 |
| Adversarial Method |  | SNR(dB) | 41.60 | 40.56 | 35.27 | 34.99 | 32.89 | 41.73 | 32.89 | 34.05 | 36.33 | 35.58 | 36.29 |
|  |  | RMSE | 0.0027 | 0.0031 | 0.0058 | 0.0053 | 0.0068 | 0.0027 | 0.0074 | 0.0062 | 0.0048 | 0.0079 | 0.0053 |
| CGAN |  | SNR(dB) | 40.09 | 40.29 | 41.18 | 41.79 | 40.72 | 39.52 | 42.63 | 40.71 | 40.87 | 43.15 | 41.10 |
|  |  | RMSE | 0.0021 | 0.0027 | 0.0033 | 0.0024 | 0.0023 | 0.0022 | 0.0026 | 0.0027 | 0.0025 | 0.0031 | 0.0026 |
| Cardio-NAFNET (proposed) |  | SNR(dB) | 54.25 | 50.20 | 51.37 | 48.87 | 51.56 | 55.23 | 49.96 | 49.25 | 52.65 | 52.04 | 51.54 |
|  |  | RMSE | 0.0016 | 0.0027 | 0.0045 | 0.0094 | 0.0012 | 0.0009 | 0.0040 | 0.0051 | 0.0021 | 0.0015 | 0.0033 |

**Table 5:**
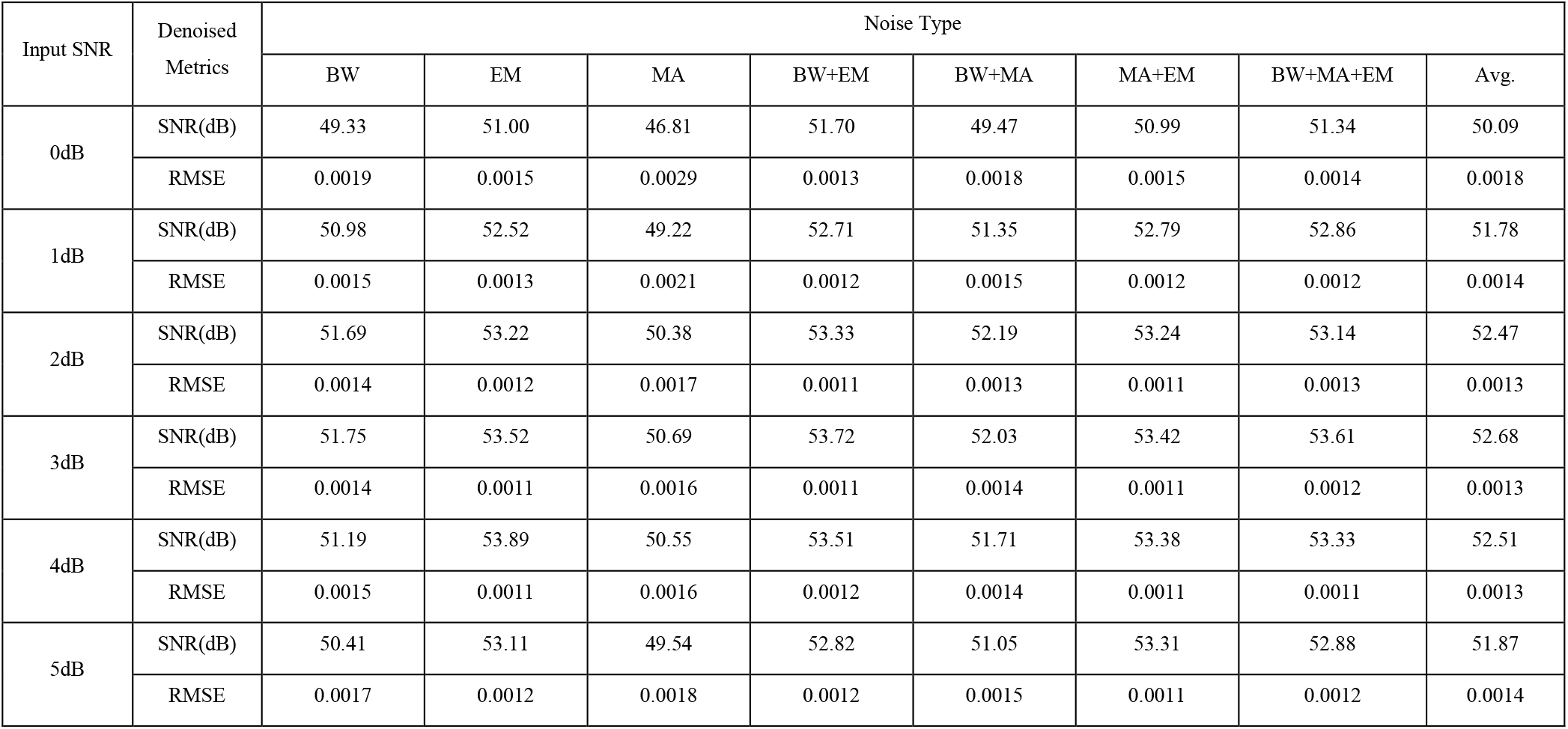
Denoising results of 8 secs samples with 64Hz sampling rate by noise type and SNR

**Table 6:**
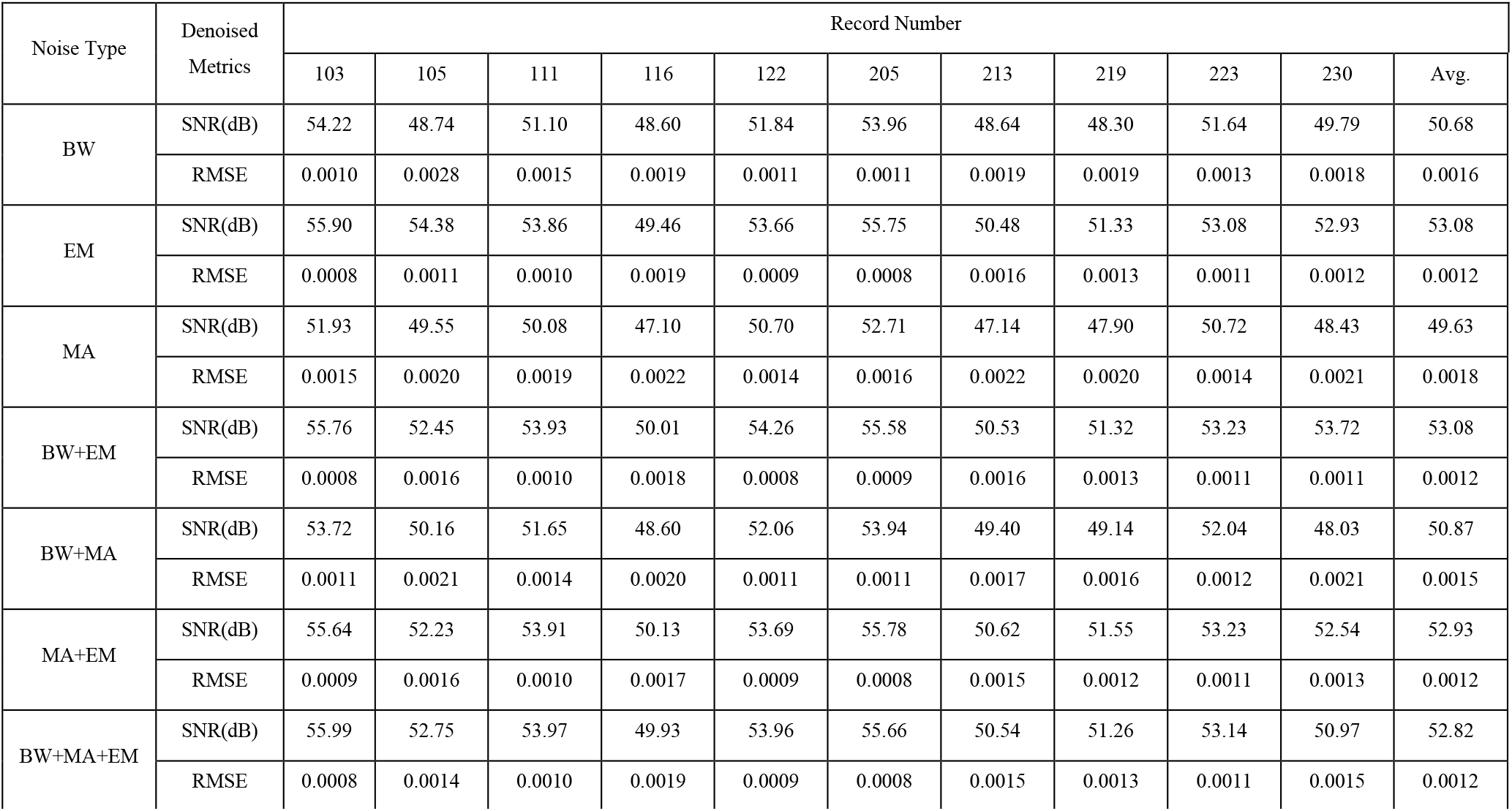
Denoising results of 8 secs samples with 64Hz sampling rate by record number and noise type

**Table 7:** Classification comparison between clean, noisy, and denoised signals

|  | Clean | Noisy | Denoised | Improved |
| --- | --- | --- | --- | --- |
| BW | 98.03% | 73.27% | 97.90% | 24.62% |
| EM | 98.02% | 71.42% | 97.88% | 26.46% |
| MA | 97.91% | 73.10% | 97.52% | 24.42% |
| BW+EM | 98.05% | 70.25% | 97.97% | 27.72% |
| BW+MA | 98.04% | 73.36% | 97.93% | 24.57% |
| MA+EM | 97.76% | 70.28% | 97.58% | 27.30% |
| BW+MA+EM | 97.83% | 69.73% | 97.83% | 27.99% |

**Table 8.**
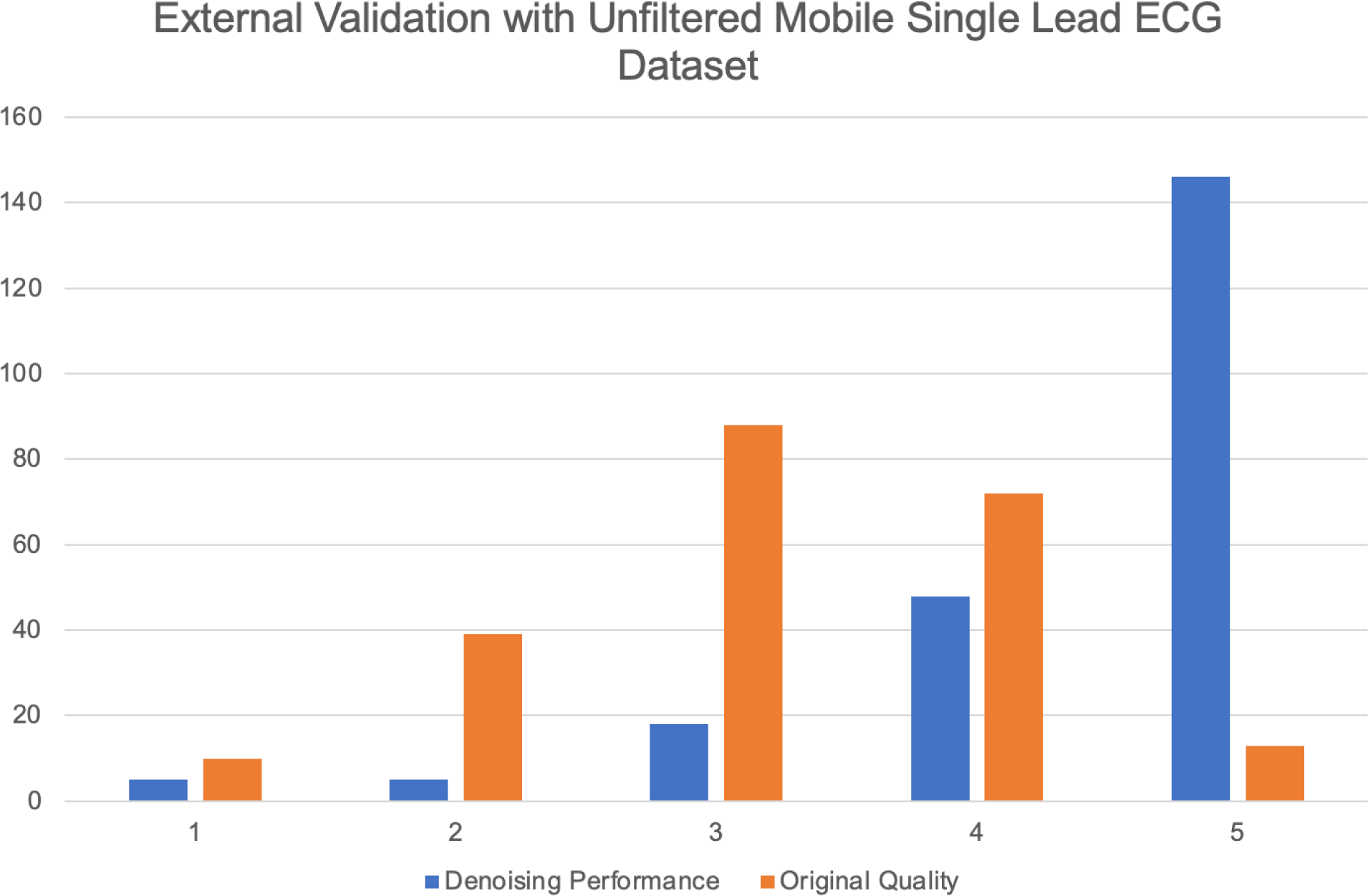
The histogram demonstrates the distribution of signal quality when the unfiltered mobile ECG samples and their corresponding denoised outputs from Cardio-NAFNet when reviewed by physicians. While the majority of the signals that were uninterpretable stayed uninterpretable, Cardio-NAFNet was able to significantly improve the quality of the signals as shown in figure 3.

## Discussion

In this work, we present Cardio-NAFNet that outperforms existing state-of-the-art methods in quantitative measures. We also augment the conventional experiment design of quantitative ECG denoising performance evaluation by qualitative evaluation methods and an external validation of mobile ECG signals for generalizability. SNR served as a popular metric to evaluate the quality of a signal or image samples, but we believe that the most critical piece of ECG denoising is not to generate signals that are just visibly good, but to enhance the clinical interpretability of the signals. Previous literature demonstrated the average SNR and the classification results of the model’s denoised outputs when the ECG records are broken down into nearly a single second[20, 21, 30, 31], containing one to two beats. While Wang et al created an extensive design to test CAE-CGAN, we believe that the model performance should be also evaluated with longer signals as the irregularities in rhythm that cannot be captured in a single beat can have significant clinical implications. Also, generalizability has been regularly concerned in numerous publications when it comes to the AI models used in medicine[29, 32, 33]. AI models within ECG domain are no exception as different device types and patient population can cause AI models to underperform when it is exposed to an unseen dataset. Demonstrating the generalizability of denoising models with mobile ECGs has been a difficult task due to a limited number of datasets with clean ECG samples and noise samples that are publicly available. Since the rise of consumer level ECG devices such as AliveCor Kardia or Apple Watch, the validation of AI-based ECG model’s generalizability with single lead mobile ECG signals has been imperative. Numerous authors have addressed the generalizability of their models by stratifying the dataset at a patient level and providing unseen leads to the model during tests using MIT-BIH Physionet’s Arrhythmia database [5-10, 12-14, 17-21, 30, 31, 34-37]. Despite the attempt, these models prove its generalizability within the Arrhythmia database, which contains Holter recordings from 48 patients recorded at a single lab during 1975 to 1979. We argue that previously suggested experimental framework does not suffice to prove the model’s generalizability, especially when the large demands arise from mobile ECG signals.

Cardio-NAFNet, with three experiments, validated its performance and addressed all the limitations above. The three experiments were designed with the following objectives:

1. Confirm superior performance in an identical training and testing environment to the current SOTA model.
2. Validate Cardio-NAFNet’s capabilities with 8 second recordings with SNR and enhanced classification results.
3. Prove the generalizability of Cardio-NAFNet through an external validation.

Our external validation highlights Cardio-NAFNet’s generalizability not only at the device level, but also at a patient population level as the data was collected from 44 sites around the world. We also note that most of the original samples that were hardly interpretable stayed uninterpretable after denoising, which is reasonable.

## Conclusion

In this paper, we propose a novel AI ECG denoising method based on NAFNet architecture and extensive experiment designs to evaluate the denoised signal’s clinical interpretability and generalizability. Cardio-NAFNet further contributes to ECG denoising where the previous methods have been limited by employing the structure of simplified attention blocks in a U-Net architecture with loss functions tailored to ECG denoising. Cardio-NAFNet consistently achieved SNR above 50dB in majority of the extensive testing environment, which is a mark that no other model in literature has achieved so far. The ECG denoising performance was not only evaluated by SNR, but also qualitatively with a pre-trained ECG classification model and expert physicians to demonstrate improved classification results and enhanced signal quality. Overall, Cardio-NAFNet shows promising results in ECG denoising in both Holter recordings and mobile single lead recordings, proving its generalizability and clinical significance.

Overall, Cardio-NAFNet provides a denoising method to standardize and stabilize the ECG recordings from mobile devices. In our future studies, we plan to apply Cardio-NAFNet to the mobile ECG data and translate innovative AI works that have been done with 12-lead ECG signals in clinics or labs to be applicable to the mobile ECG recordings.

## Data Availability

The data used to train the model (Physionet MIT-BIH Arrhythmia Database and Noise-Stress Database) are public on Physionet.

https://physionet.org/

